# COVID-19: How Many Years of Life Lost?

**DOI:** 10.1101/2020.06.08.20050559

**Authors:** Harry P. Wetzler, Erica A. Wetzler, Herbert W. Cobb

## Abstract

**Background:** COVID-19 was the leading cause of death in the United States over the three-month period March through May 2020. Another perspective is COVID-19’s toll in terms of years of life lost. We calculated years of life lost for COVID-19 and other leading causes of death over those three months in the US. We also predicted years of life lost for COVID-19 and ischemic heart diseases (which includes heart attacks) for March through August 2020.

**Methods:** Years of life lost are the sum of differences between life expectancy at age of death and age at death. Average years of life lost, years of life lost divided by the number of deaths, were also calculated. We used the COVID-19 Projections Using Machine Learning model to predict years of life lost from COVID-19 through the end of August 2020.

**Results:** COVID-19 caused 12,035 more deaths than ischemic heart diseases during March through May 2020 but ischemic heart diseases years of life lost were 1.5% greater than those for COVID-19. Average years of life lost were 10.8 and 12.4 for COVID-19 and ischemic heart diseases, respectively. At the end of August, COVID-19 may overtake ischemic heart diseases as the leading cause of deaths and years of life lost in the US.

**Conclusion:** Each COVID-19 death causes more than a decade of lost life in the US. We are reminded of a Danish Proverb that states “Prediction is difficult, especially when dealing with the future.” We suggest that while dying is bad, losing life is even worse.

## Introduction

During the months of March, April, and May of 2020, COVID-19 became the number one killer in the United States (US).^1^ Not since the great influenza pandemic of 1918 has there been a contagion with such a nationwide impact.^2^ The US recorded its 100,000^th^ COVID-19 death on May 27. Although most models predict a slowing in the death increase in the coming months and the re-emergence of heart problems as the mortality leader, COVID-19 could be the number two cause of death for all of 2020, after ischemic heart diseases (IHD). Moreover, Osterholm suggested that we are in the early phase of the pandemic, and there could be 800,000 COVID-19 deaths in the US over the next 18 months.^3^

In 1947 Dempsey stated that deaths do not tell the entire story regarding the seriousness of a disease, and she introduced the concept of “potential years of life lost”.^4^ A year later Greville stated the justification in simple terms “from the standpoint of society, the death of a person in the prime of life constitutes a greater loss than the death of one of advanced years.”^5^ Since then years of life lost (YLLs) have been the subject of considerable research perhaps culminating in the Global Burden of Disease studies.^6,7,8^ Furthermore, the initial metric, years of life lost, has been modified to include disability-adjusted life years (DALYs) and years lived with disability (YLDs) in an attempt to account for the fact that not all life years are equal due disability.

This study had two goals: tally deaths and YLLs for major causes during March through May 2020 (January through March 2020 for influenza and pneumonia due to seasonality) and calculate YLLs for IHD during March through August 2018 and compare them to those projected for COVID-19 over the same six months in 2020.

## Methods

We used the Centers for Disease Control and Prevention (CDC) WONDER Multiple Cause of Death Online Database, which includes mortality estimates for the United States stratified by age, sex, and cause of death.^9^ The latest year in the WONDER Database is 2018. Only deaths from March through May 2018 (January through March for influenza and pneumonia due to seasonality) were used for comparison to COVID-19 deaths because, except for 2020, aggregate causes of death are quite stable from year to year. The maximum year to year change for 2014-2018 was 11.5% (Nontransport accidents 2015-2016) and the minimum was −3.1% (Malignant neoplasms of trachea, bronchus and lung 2015-2016). Total COVID-19 deaths were obtained from the Johns Hopkins database and the age strata posted by CDC were used.^1,10^

Since COVID-19 is basically a single cause of death, we used more discrete categories for comparison instead of the broad ones such as Diseases of Heart or Malignant Neoplasms that are customary. We also included influenza and pneumonia because early on some policy-makers claimed the coronavirus was no worse than the flu.^11^ The categories used and their UCD - ICD-10 Codes are listed below.

### Cause

- COVID-19 (U071)
- Ischemic Heart Diseases (IHD) (I20-I25)
- Other Heart Diseases (HD) (I26-I51)
- Chronic Lower Respiratory Diseases (CLR) (J40-J47)
- Cerebrovascular Diseases (CVD) (I60-I69)
- Cancer of Trachea, Bronchus and Lung (C33-C34)
- Nontransport Accidents (W00-X59, Y86)
- Influenza and Pneumonia (J09-J18)
- Cancer of Breast (C50)
- Transport Accidents (V01-V99, Y85)
- Cancer of Prostate (C61)

YLLs were calculated using 11 age groups at the time of death: less than 1 year, 1 to 4 years, 5 to 14 years, 15 to 24 years, 25 to 34 years, 35 to 44 years, 45 to 54 years, 55 to 64 years, 65 to 74 years, 75 to 84 years, and 85 years or older. These age groups are consistent across the WONDER Database and the posted COVD-19 deaths by age group.^10^ Life expectancy data were obtained from the US Life Table for 2017.^12^ YLLs are the sum of differences between the mid-points in each age stratum and life expectancy at that age. To facilitate comparisons outside the US, we also calculated YLLs using Standard Expected YLLs (SEYLLs) based on the World Health Organization (WHO) Standard Life Table for Years of Life Lost.^13^ The WHO Standard Life Table has life expectancies projected through year 2050 that are greater than those in the 2017 US life table. Average years of life lost per decedent are the total YLLs divided by the number of decedents. In this preliminary study we did not calculate DALYs or adjust YLLs for quality of life or comorbidity.^8,14^

The COVID-19 Projections Using Machine Learning model is a susceptible, infectious, and recovered (or deceased) (SEIR) model that uses artificial intelligence to minimize the error between the projected outputs and the actual results.^15^ The model’s 95% confidence interval projections made on May 31 for August 31 COVID-19 deaths were used with the COVID-19 decedent age distribution posted by CDC to estimate COVID-19 YLLs through August 31, 2020 and compared to those for Ischemic Heart Diseases for March through August 2018.

We used Microsoft Excel for all calculations.

## Results

A total of 710,702 deaths occurred in the US during the three-month period March through May 2018. Deaths by major underlying cause during that time along with COVID-19 deaths in March through May 2020 and influenza and pneumonia deaths in January through March 2018 are listed in Table 1. Table 1 also shows the YLLs, average years of life lost, and percent of total YLLs for each cause.

**Table 1.** United States Mortality, Years of Life Lost, and Average Years of Life Lost by Cause of Death (March - May 2018).

| **Cause of Death** | **Total Deaths** | **Years of Life Lost** | **Average Years Lost** | **Percent of Total YLLs** |
| --- | --- | --- | --- | --- |
| **COVID-19\*** | 104,381 | 1,128,186 | 10.8 | 10.5% |
| **Ischemic Heart Diseases** | 92,346 | 1,144,811 | 12.4 | 10.7% |
| **Other Heart Diseases** | 57,580 | 662,543 | 11.5 | 6.2% |
| **Chronic Lower Respiratory Diseases** | 42,068 | 503,304 | 12.0 | 4.7% |
| **Cerebrovascular Diseases** | 37,051 | 400,330 | 10.8 | 3.7% |
| **Cancer of Trachea, Bronchus and Lung** | 35,693 | 527,690 | 14.8 | 4.9% |
| **Nontransport Accidents** | 31,117 | 858,069 | 27.6 | 8.0% |
| **Influenza and Pneumonia\*\*** | 26,850 | 326,920 | 12.2 | 3.0% |
| **Cancer of Breast** | 10,641 | 193,186 | 18.2 | 1.8% |
| **Transport Accidents** | 10,103 | 373,047 | 36.9 | 3.5% |
| **Cancer of Prostate** | 7,795 | 77,138 | 9.9 | 0.7% |
| **All Causes** | 710,702 | 10,739,230 | 15.1 | 100% |

\*COVID-19 deaths include those in March through May 2020
\*\*Influenza and Pneumonia deaths include those in January through March 2018

COVID-19 is the leading cause of death but YLLs for ischemic heart diseases are 1.5% higher. Average years of life lost are equal for COVID-19 and cerebrovascular diseases with older decedents in those two categories. Conversely, the average years of life lost are markedly higher for both categories of accidents indicating relatively more deaths among young people. In these data the correlation coefficient between working years of life lost, years lost before age 65, and average years of life lost is 0.95. COVID-19 and ischemic heart diseases together account for over a fifth of the total YLLs.

Figure 1 illustrates the relationships between numbers of deaths and YLLs for the major causes of death.

**Figure 1.**
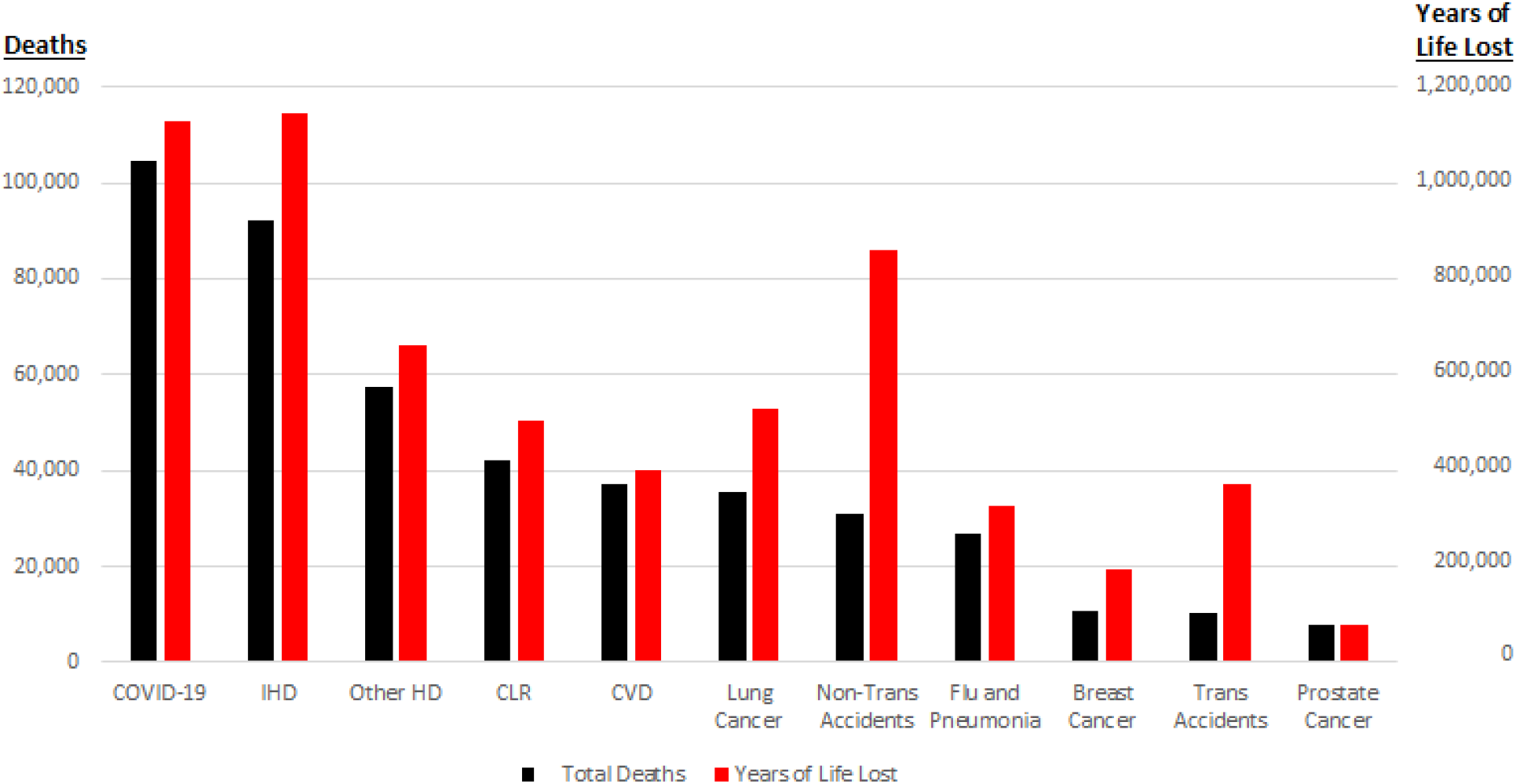
Deaths and Years of Life Lost (March-May 2020)

Although the WHO SEYLLs are greater than YLLs calculated using the US life table, the order of SEYLLs by cause of death is identical to that for US-based YLLs, and the COVID-19 and IHD SEYLLs are essentially equal (1,685,399 and 1,686,091 respectively). The SEYLLs order of average years of life lost is also identical to that for US-based YLLs.

On May 31, 2020, the 95% confidence interval for cumulative deaths on August 31, 2020 from the COVID-19 Projections Using Machine Learning model was 127,736 to 335,902.^15^ YLLs for those numbers of deaths are 1,380,615 and 3,630,546, respectively. From March through August 2018 there were 177,300 IHD deaths and 2,221,253 YLLs. If COVID-19 deaths total 205,513 or more on August 31, then COVID-19 YLLs will equal or exceed those for IHD over the 6-month period.

## Discussion

COVID-19 killed more than 100,000 people in the US in less than 90 days. Although we do not know the extent to which COVID-19 is displacing other deaths, we do know that most of these were excess deaths that would have taken about 21 months to occur in the absence of the virus.^16^,^17^ And, as Mary Dempsey wrote in 1947, deaths fail to tell the entire story.^4^ Age at death matters and YLLs measure when deaths occur. IHD has been the leading cause of YLLs in the US since 1990.^18^ For at least three months, March through May 2020, COVID-19 was not only the leading cause of death but it rivaled IHD for the lead in YLLs. Our findings echo those of Hanlon, et al, who found that in the United Kingdom female COVID-19 decedents lost an average of 12 years of life with 14 for men compared to 10.8 years in the US.^19^ Moreover, these results are not based on an arcane model; they reflect the intuitive concept of dying too soon.

We need to be clear about the purpose of modelling. Models are never meant to provide exact numbers. Rather, by providing a range or a window of estimates for outcomes, including deaths, models serve the important purpose of guiding prevention and control measures as well as informing policy and planning regarding the disease modeled. Indeed, the COVID-19 Projections Using Machine Learning model makes a point estimate of the cumulative number of deaths by August 31 and that estimate changes slightly every day. That does not make the model wrong given its purpose; we use the 95% confidence interval of what is likely to occur. Three months of excess deaths and YLLs are bad enough but the model suggests there is a 33% chance of over 200,000 COVID-19 deaths in the US by the end of August.^15^ Thus, there is about a 30% chance that by the end of August COVID-19 YLLs will equal or exceed those for IHD over the previous six months. Although we anticipate that IHD YLLs will be the leader for all of 2020, there may be a second COVID-19 wave and we are reminded of Osterholm’s forecast that there could be 800,000 COVID-19 deaths in the US by the end of 2022.^3^

Several factors may influence our YLL estimates and comparisons. The first factor is the accuracy of underlying cause of death numbers. The tallies of COVID-19 deaths have been criticized for under-reporting. For example, the government counts 26,000 COVID-19 deaths in nursing homes. But because the federal government is not requiring nursing homes to report deaths prior to May, the under-reporting of deaths at nursing homes could be in the thousands.^20,21^ On the other hand, there is reason to question whether COVID-19 deaths are being over-counted because of higher payment for COVID-19 diagnosis in Medicare recipients.^22^ In comparison, WHO reports that the relative uncertainty for deaths from IHD is about ±12% for high-income countries.^23^ The second factor is using 2018 data for causes of death other than COVID-19. Although year to year cause of death counts are quite stable, there may be some displacement of deaths in 2020. We will not be able to accurately assess displacement or other changes for six months or more. The third factor is variation in life expectancy which is a random variable but with small standard errors (e.g., approximately 0.04 years at birth in 2017).^24^ The final factor is assumed stable age distributions of COVID-19 deaths.

Regarding the accuracy of the data, we do not know the exact numbers for any cause of death. For example, influenza deaths in the US are estimated each year.^25^ Cumulative COVID-19 deaths on May 31, 2020 ranged from 98,600 to 106,432 on two prominent tracking sites.^26,27^ The Global Burden of Disease Study reported 544,800 IHD deaths in the US in 2016 whereas CDC WONDER included only 363,452—33% less.^18,9^ Yet to dismiss counts, analyses, and models because they lack exact accuracy is a mistake and undermines their purpose. We are well advised to pay attention to Spiegelhalter who wrote “my cold, statistical approach is to wait until the end of the year, and the years after that, when we can count the excess deaths.”^28^ However, it is not merely an accounting exercise to understand the severity of the current health crisis even if such models are not totally accurate. Timely evidence-based analysis should support institutional policy-making and personal decision-making else those decisions are, by definition, ill-informed. We acknowledge the limitations of this analysis, and hence, will update it in September after the August mortality counts stabilize. We will also include quality of life and comorbidity adjusted YLLs.

## Conclusions

COVID-19 killed more than 100,000 people in the US in less than 90 days and most of these were excess deaths that would have taken about 21 months to occur without the virus. For at least three months, March through May 2020, COVID-19 was not only the number one cause of death, but it was number two only to ischemic heart diseases in years of life lost. If there is a significant second COVID-19 wave in the fall, 2020 will be the year ischemic heart diseases are dethroned as the number one killer as well as the number one cause of years of life lost since 1990 in the USA. In this context we are reminded of a Danish Proverb that states “Prediction is difficult, especially when dealing with the future.”^29^ We suggest that while dying is bad, losing life is even worse.

## Data Availability

All data used in the study are available online from the sources indicated in the manuscript.

## References

1. Time_series_covid19_deaths_US.Csv., 2020. https://github.com/CSSEGISandData/COVID-19/tree/master/csse_covid_19_data/csse_covid_19_time_series Accessed June 2, 2020

2. Barry J. The Great Influenza: The Epic Story of the Deadliest Plague in History., 2004. doi:10.1080/00396338.2004.9688606

3. Bergen P. Michael Osterholm: Infectious disease expert says we’re only in the second inning of the pandemic - CNN. published April 21, 2020. https://www.cnn.com/2020/04/21/opinions/bergen-osterholm-interview-two-opinion/index.html

4. Dempsey M. Decline in Tuberculosis The Death Rate Fails to Tell the Entire Story. Am Rev Tuberc. 1947;56(2):157–164.

5. Greville TN. Comments on Mary Dempsey’s Article on “Decline in Tuberculosis: The Death Rate Fails to Tell the Entire Story.” Am Rev Tuberc. 1948;57(4):417–419. doi:10.1164/art.1948.57.4.417

6. Wetzler HP. Loss of Working Years in Accidents. N Engl J Med. 1976;294(24). doi:10.1056/NEJM197606102942417

7. Premature Mortality in the United States: Public Health Issues in the Use of Years of Potential Life Lost. https://www.cdc.gov/mmwr/preview/mmwrhtml/00001773.htm

8. Global Burden of Disease Study 2017 (GBD 2017) Cause-Specific Mortality 1980-2017 | GHDx. http://ghdx.healthdata.org/record/ihme-data/gbd-2017-cause-specific-mortality-1980-2017 Accessed June 1, 2020.

9. Multiple Cause of Death Data on CDC WONDER. https://wonder.cdc.gov/mcd.html Accessed June 1, 2020.

10. Provisional Death Counts for Coronavirus Disease (COVID-19). https://www.cdc.gov/nchs/nvss/vsrr/covid19/index.htm Accessed June 2, 2020.

11. Faust J. Comparing COVID-19 Deaths to Flu Deaths Is like Comparing Apples to Oranges - Scientific American Blog Network. https://blogs.scientificamerican.com/observations/comparing-covid-19-deaths-to-flu-deaths-is-like-comparing-apples-to-oranges/

12. Arias E, Xu J. United States Life Tables, 2017. NVSR Volume 68, Number 7. National Vital Statistics Report., 2017. https://www.cdc.gov/nchs/products/index.htm.

13. World Health Organization. Global desease estimates, 2000-2016. 2018;(June). http://www.who.int/healthinfo/global_burden_disease/GHE2016_DALY_Global_2000_2016_.xls?ua=1

14. Moving Beyond “Lives-Saved” From COVID-19 | Avalon Health Economics LLC. https://avalonecon.com/moving-beyond-lives-saved-from-covid-19/ Accessed June 6, 2020.

15. COVID-19 Projections Using Machine Learning. https://covid19-projections.com/about/ Accessed May 31, 2020.

16. Estimating the early death toll of COVID-19 in the United States. https://weinbergerlab.github.io/excess_pi_covid/ Accessed June 4, 2020.

17. Wetzler HP, Wetzler EA. Covid-19 excess deaths in the United States through July 2020. medRxiv. published online May 19, 2020:2020.04.02.20051532. doi:10.1101/2020.04.02.20051532

18. Murray CJL, Mokdad AH, Ballestros K, et al. The state of US health, 1990-2016: Burden of diseases, injuries, and risk factors among US states. JAMA - J Am Med Assoc. 2018;319(14):1444–1472. doi:10.1001/jama.2018.0158

19. Hanlon P, Chadwick F, Shah A, et al. COVID-19 – exploring the implications of long-term condition type and extent of multimorbidity on years of life lost: a modelling study. Wellcome Open Res. 2020;5(May):75. doi:10.12688/wellcomeopenres.15849.1

20. Centers for Medicaid and Medicare Services. Interim Final Rule Updating Requirements for Notification of Confirmed and Suspected COVID-19 Cases Among Residents and Staff in Nursing Homes., 2020. https://nhsn.cdc.gov/RegistrationForm/index Accessed June 4, 2020.

21. The government counts 26,000 COVID-19 deaths in nursing homes. That’s at least 14,000 deaths too low. published June 2, 2020. https://www.nbcnews.com/health/health-news/government-counts-26-000-covid-19-deaths-nursing-homes-s-n1221496

22. Miltimore J. Physicians Say Hospitals Are Pressuring ER Docs to List COVID-19 on Death Certificates. Here’s Why - Foundation for Economic Education. published April 29, 2020. https://fee.org/articles/physicians-say-hospitals-are-pressuring-er-docs-to-list-covid-19-on-death-certificates-here-s-why/

23. Years of Life Lost (Percentage of Total). https://www.who.int/whosis/whostat2006YearsOfLifeLost.pdf?ua=1

24. Li N, Tuljapurkar S. On the accuracy of life expectancy. Popul Assoc Am 2012, Annu Meet Progr. published online 2012:1-14. http://paa2012.princeton.edu/papers/120176

25. CDC. Weekly US. Influenza Surveillance Report (FluView). Centers for Disease Control and Prevention. Publ March. 2020;6:2020. https://www.cdc.gov/flu/weekly/#S2 Accessed June 2, 2020.

26. US Historical Data | The COVID Tracking Project. https://covidtracking.com/data/us-daily Accessed June 1, 2020.

27. United States Coronavirus - Worldometer. https://www.worldometers.info/coronavirus/country/us/ Accessed June 1, 2020.

28. Spiegelhalter D. Coronavirus deaths: how does Britain compare with other countries? The Guardian. published April 30, 2020. https://www.theguardian.com/commentisfree/2020/apr/30/coronavirus-deaths-how-does-britain-compare-with-other-countries

29. It’s Difficult to Make Predictions, Especially About the Future – Quote Investigator. https://quoteinvestigator.com/2013/10/20/no-predict/ Accessed June 4, 2020.

